# AI-enabled conversational agent increases engagement with cognitive-behavioral therapy: A randomized controlled trial

**DOI:** 10.1101/2024.11.01.24316565

**Authors:** Jessica McFadyen, Johanna Habicht, Larisa-Maria Dina, Ross Harper, Tobias U. Hauser, Max Rollwage

**Author notes:** **ClinicalTrials.gov ID:** NCT06459128.

## Abstract

Timely support after referral to mental healthcare is crucial, yet patients often face prolonged wait times without intervention. Digital mental health interventions offer scalable solutions, but many struggle to achieve acceptable patient engagement. Tailoring and personalizing materials to individual needs is paramount for driving engagement, a task that generative artificial intelligence AI (genAI) is potentially able to achieve. To examine this promise, we conducted a randomized controlled trial using a genAI-enabled therapy app, Limbic Care, which delivers personalized cognitive behavioral therapy (CBT) materials, against PDF workbooks delivering static CBT content, as commonly used in standard care. Adults with elevated symptoms of anxiety or depression (N = 540) were randomly assigned to the app or control group for six weeks. The app group exhibited a threefold increase in engagement (2.4 times higher usage frequency, 3.8 times longer usage durations). While both groups showed similar overall symptom improvement, participants who engaged with the app’s clinical personalization capabilities experienced significantly greater reductions in anxiety symptoms and enhanced well-being than those who engaged with the standard CBT materials. Importantly, the app was safe, with no increase in adverse events compared to standard care. Our findings suggest that genAI-enabled therapy apps can safely enhance patient engagement and improve clinical outcomes through clinically personalized interventions.

## Introduction

Without timely intervention, symptoms of mental ill health can progressively worsen (Anderson et al., 2019). Prolonged waiting periods without support can exacerbate symptoms and elevate the risk of adverse outcomes and drop-out from treatment (Peipert et al., 2022; Punton et al., 2022). Ensuring that patients receive support and begin engaging with their treatment during this precarious waiting period is a key contributing factor to positive patient outcomes (Peipert et al., 2022). Similarly, engaging with therapeutic material in-between therapy sessions (i.e., “homework”) has been shown to markedly improve clinical outcomes and therapy adherence (Mausbach et al., 2010). Especially for cognitive-behavioral therapy (CBT), there is evidence that homework encourages patients to incorporate what they have learned from therapy into their everyday life, reinforcing new skills, promoting behavioral and cognitive change, and eventually leading to better treatment outcomes (Prasko et al., 2022).

Encouraging consistent engagement with therapeutic activities – either in-between therapy sessions or while on a waitlist for treatment – remains a significant challenge. Limited availability of clinical staff and the high costs of continuous human supervision restrict widespread access to personalized care through crisis hotlines, teletherapy, or messaging services (Tran, 2024). Additionally, patients often struggle to engage with generic, “one-size-fits-all” content designed to cater to a broad audience (Oewel et al., 2024). Both traditional methods (e.g., physical workbooks) and current digital solutions (e.g., static apps or online platforms) do not overcome these barriers, as they lack the personalized, interactive support needed to keep patients engaged and help them make meaningful progress due to their static content (Gilbody et al., 2015; Malik et al., 2022; Torous et al., 2018).

The advent of large language models (LLMs) introduces a transformative opportunity to overcome these limitations. Unlike traditional digital interventions, LLM-powered generative AI (genAI) can facilitate highly interactive and personalized experiences that closely mimic therapist-patient interactions (Kuhail et al., 2024; Meyer & Elsweiler, 2024). By providing dynamic, responsive, and tailored support, LLM-powered applications can adapt in real-time to each user’s unique context, effectively bridging the gap when an actual therapist is not available (Rollwage, Habicht, Juchems, et al., 2024; Rollwage, Habicht, Juechems, et al., 2024). This level of personalization and engagement is unique to genAI and is not attainable with standard digital interventions. GenAI-enabled solutions therefore offer a critical advancement in enhancing patient engagement and clinical outcomes – provided this can be done safely and remains clinically relevant and effective (Cabrera et al., 2023).

To address the challenges with therapeutic engagement, we leveraged these recent innovations in genAI and developed a clinically validated, genAI-powered smartphone application called Limbic Care. This app features a conversational chatbot designed to deliver standard CBT materials and is intended for use as a first-line, evidence-based treatment for depression and anxiety. The integration of genAI aims to provide a personalized, user-centric experience that cultivates a relationship between the user and the application, which has the potential to increase engagement both in terms of quantity (e.g., how often the tool is used and for how long) and quality (e.g., enabling more meaningful interactions with material tailored to the user’s personal problems).

The aim of this randomized controlled trial (RCT) was to evaluate the effectiveness of this evidence-based, genAI-powered app in improving engagement and symptoms of anxiety and depression in adults with mild to severe depression or anxiety symptoms. We focused specifically on adults who were not currently waiting for or undergoing therapy for their mental health, and thus there was no human therapist in the loop. As an active control, we compared the intervention group to a control group who received a standard PDF workbook used in typical care settings.

Our primary objectives were to evaluate: 1) whether a genAI-enabled app increases patient engagement with therapeutic activities, 2) its effectiveness in reducing symptoms of anxiety, depression, and sleep disturbances, and improving overall well-being, and 3) how it compares in safety to the standard of care. We hypothesized that the genAI-powered app would be at least as effective as (i.e., non-inferior to) the standard PDF workbook in reducing symptoms, while providing the additional benefit of enhanced patient engagement through its interactive and personalized features. By systematically assessing these outcomes, this study sought to provide empirical evidence on the potential of LLMs to maintain clinical effectiveness while enhancing patient engagement in digital mental health interventions, benchmarked against standard care.

## Methods

### Study design

This was a 6-week, two-arm, parallel-group RCT comparing the effectiveness of a genAI-enabled digital CBT app (Limbic) in delivering CBT exercises to a static PDF workbook equivalent. The study was conducted as an open-label trial due to the nature of the interventions. Participants were randomly allocated to the intervention group (Limbic app) or control group (PDF format) in a 3:2 ratio, in order to adequately power finer-grained behavioral subgrouping based on app engagement patterns. Ethical approval was obtained from the UCL Research Ethics Committee [6218/003]. The study design, including primary and secondary and additional outcome measures and their analyses, was preregistered on the ClinicalTrials.gov registry (NCT06459128) prior to the commencement of data collection. All procedures were conducted in accordance with the Declaration of Helsinki.

### Participants

Eligible participants were United States residents aged 18+ years with anxiety and/or depression symptoms above a clinical threshold, as defined by the Generalized Anxiety Disorder 7-item questionnaire (GAD-7 scores ≥ 7) and the Patient Health Questionnaire (PHQ-9; scores ≥ 9), respectively. Inclusion criteria included fluency in English, access to a smartphone, and not currently receiving psychological therapy from a mental health professional. Exclusion criteria included high alcohol intake (≥ 10 alcohol units per week), frequent recreational drug usage (more than weekly), recent changes in dosage or frequency of prescription medication for mental health (in the last 8 weeks), previous usage of the Limbic app, and self-reporting being at risk of self-harm or causing harm to others. All participants were recruited online via the Prolific platform (https://www.prolific.com/) and provided informed consent.

### Intervention

#### INTERVENTION: LIMBIC CARE

Limbic Care is a smartphone-based application designed to deliver therapeutic content and support to adults aged 18 years or older in settings outside in-person therapy, such as between therapy sessions or while waiting to begin treatment. The intervention is centered around an AI-powered conversational chatbot that uses large language models (LLMs) and clinically-specialized machine learning (ML) algorithms to assist users in completing therapeutic exercises (psychoeducation lessons and CBT activities), provide emotional support (through the “Let’s Chat” feature), and guide users through structured problem exploration sessions and clinically-personalized exercise suggestions (referred to as “guided sessions”). The primary aim of the app is to facilitate self-directed engagement with therapeutic materials, enabling patients to independently apply clinically-validated techniques in their daily lives, whether between therapy sessions or while on a waitlist. The app therefore serves to enhance accessibility and practical application of CBT principles.

The app offers three main functionalities:

1. **“Let’s Chat” for Emotional Support**: The “Let’s Chat” feature allows users to engage in free-flowing conversation to express their emotions and discuss challenges they are facing. The chatbot provides active listening and empathetic responses, following principles of person-centered therapy to offer emotional support. Thus, this component is intended to create a non-judgmental space where users can “vent” or talk through problems in a supportive environment.
2. **Delivery of CBT Materials and Exercises (Activities and Psychoeducation)**: Participants have access to a range of clinically-validated CBT content delivered through conversational interactions with the chatbot. There are two primary types of materials provided:

○ *Psychoeducation*: Short, digestible content focused on educating users about specific psychological concepts or coping strategies (e.g., understanding cognitive distortions, building resilience).
○ *CBT Activities*: Structured exercises inspired by CBT principles, such as thought records, behavioral activation tasks, and mindfulness practices. These exercises are designed to help users actively work through negative thoughts and maladaptive behaviors.
3. **Guided Sessions**: This feature allows users to explore a particular issue or challenge they are experiencing. Through guided questioning, the chatbot and a network of clinically-validated ML algorithms identifies core patterns or mechanisms associated with the user’s problem. Based on this exploration, the underlying ML architecture then recommends tailored CBT exercises that address the identified issues, providing a personalized therapeutic experience.

Participants could select one of three therapeutic “courses” offered by the app, targeting specific psychological concerns for managing sleep problems, worry, or low mood. For each course, different interventions and psychoeducational materials were made available in the app’s “to-do list” section according to a predefined schedule. This helps ensure that users receive a structured sequence of activities tailored to their primary concern, delivered over time to support incremental progress.

#### CONTROL: PDF WORKBOOK

Participants in the control group received access to a digital PDF workbook containing therapeutic content identical to the psychoeducation and CBT interventions delivered by the Limbic Care app. Participants were provided with a unique website URL to access the workbook, which was presented as a static, non-interactive PDF. The website required users to enter their unique participant ID to view the PDF, allowing us to track the viewing behavior of participants. Thus, this implementation enabled us to quantify engagement with the control materials.

Participants could view their PDF on a smartphone or computer, and could also print it out. Similar to the app intervention, participants could choose from one of three workbooks with content tailored to managing sleep problems, worry, or low mood. Each workbook contained a combination of psychoeducational material and CBT intervention worksheets. These materials were presented as static documents, consisting of text, images, and blank response boxes. Thus, the PDF workbook served as an active comparator representing the standard delivery of CBT homework within healthcare settings. The consistency in content between the intervention and control conditions meant that any comparisons between groups would indicate effects of genAI-enabled features of the Limbic app.

### Procedure

Participants were recruited on Prolific and completed a screening questionnaire. Demographic information was retrieved from Prolific’s prescreening database. Eligible participants (see **<u>Participants</u>** section for inclusion/exclusion criteria) were invited to participate in the full study several days later. This began with a baseline questionnaire which, upon completion, automatically randomly assigned participants to either the intervention or control condition (allocation logic enabled by the Gorilla software; (Anwyl-Irvine et al., 2020)). Those in the intervention condition received instructions for installing and signing in to the Limbic app on their smartphone. Those in the control condition received a URL to access the PDF workbook of their choice (a course on sleep problems, worry, or low mood). Both interventions required participants to sign in with a unique participant identifier, allowing us to match engagement data with survey data.

Over a period of 6 weeks, participants were invited to participate in a weekly survey containing questions about mental health, app/PDF engagement, and safety (see Outcome measures), as well as quantitative and qualitative measures of user experience and feedback. Participants were compensated at a rate of £9/hr for completing the surveys and were awarded a £3 bonus at the end of the study if they completed all 6 weekly surveys. Critically, participants were not financially compensated for their engagement with either the Limbic app or PDF workbook. They were encouraged to engage with their assigned materials 4 times per week but were clearly instructed that their engagement would not affect their payment.

### Outcome measures

#### PRIMARY OUTCOMES

Our primary outcome measures included: 1) engagement with the therapeutic materials, and 2) change in anxiety and depression symptom severity. We operationalised engagement as the total time spent in the Limbic app or viewing the PDF, as well as the total number of times the app or PDF website was opened. These measures were passively collected by tracking functionality included in the app and in the host website for the PDF workbooks. We excluded participants who self-reported as having printed out the PDFs at any point throughout the study, as we could not track engagement outside of the PDF hosting website. Both therapy completion and engagement duration were summed per week and analyzed over time, as well as summed across the full 6-week intervention period.

For symptom severity, we measured anxiety (GAD-7) and depression (PHQ-9) at baseline and then at weekly intervals throughout the 6-week intervention period. Both the GAD-7 and PHQ-9 scales are validated self-report scales used widely in healthcare settings as markers of mental illness (Kroenke et al., 2001; Spitzer et al., 2006). Items are rated on a 4-point Likert scale ranging from 0 (“Not at all”) to 3 (“Nearly every day”), resulting in a total score between 0 and 21 (for GAD-7) or 27 (for PHQ-9). Scores on each scale were collected each week and analyzed from baseline to the end of the intervention period, with the total change from baseline to week 6 per scale used as the overall outcome measure.

#### SECONDARY OUTCOMES

As a secondary outcome, we measured device safety by collecting self-reported incidences of adverse health events experienced by participants each week. In each weekly survey, participants were asked if they had experienced any new adverse physical or mental health events in the past week and, if so, to describe the event. Responses were analyzed to detect any “Serious Adverse Events” (SAEs), as according to ISO 14155:2020 criteria. Both the number of incidences per group and the number of unique reporting participants in each group were summed across the 6-week study period.

#### ADDITIONAL MEASURES

##### Work and Social Adjustment Scale (WSAS)

Functional impairment was assessed using the Work and Social Adjustment Scale (WSAS), a 5-item self-report measure evaluating the impact of mental health symptoms on daily functioning across work, home management, social activities, and close relationships. Each item is scored from 0 to 8, yielding a total score range of 0 to 40, with higher scores indicating greater impairment. Change in WSAS scores was analyzed from baseline to week 6, with weekly measurements.

##### Mini Sleep Questionnaire (MSQ)

Sleep quality and disturbances were evaluated using the Mini Sleep Questionnaire (MSQ), a 10-item self-report measure assessing excessive daytime sleepiness and sleep disturbances. Each item is rated on a 7-point scale, with higher scores indicating greater sleep impairment. Change in MSQ scores was analyzed from baseline to week 6, with weekly measurements.

##### User experience

Custom rating scales and free-text responses were included in the baseline and weekly questionnaires to gauge participants’ satisfaction, acceptability, and perceived effectiveness of the allocated digital tool. Each item was rated on a 7-point Likert scale, with higher scores indicating more positive user experience. Measures included ease of use (“How easy was it to navigate the app/workbook?”), usefulness (“How useful did you find the app/workbook for your mental health?”), and motivation (“How motivated were you to use the app/workbook to improve your mental health?”). A full list of measures can be found in the **<u>Supplementary Materials</u>**.

##### Beliefs and attitudes towards AI & psychotherapy

Custom rating scales and free-text responses were included in the baseline and final questionnaire in Week 6 to measure changes in beliefs and attitudes relating to digital mental health support tools. These included pre- and post-measures of trust in AI-enabled mental health tools (“In general, how much do you trust wellbeing apps that use artificial intelligence (AI)?”), preferences for workbooks vs apps for mental health support (“Imagine a mental wellbeing course was offered to you in the form of a [digital workbook (i.e., a PDF you could open on your computer or print out)]/[app that included an AI chatbot and interactive activities]. Which format would you prefer?”), and interest in pursuing therapy from a mental health professional (“How likely are you to arrange to see a therapist in the next 3 months?”).

##### Demographics

Demographic information including age, biological sex at birth, sexuality, education level, employment status, student status, and ethnicity were collected from Prolific’s prescreening database for all participants. Additional background information was collected in the baseline survey, including prior experience with therapy (experience with CBT, prior diagnosis), psychiatric medication status, how comfortable they feel with using digital tools, accessibility issues (e.g., deafness, blindness, etc.), and previous experience with mental wellbeing apps.

### Data analysis

To evaluate group differences on baseline characteristics, continuous variables were analyzed with independent samples t-tests, and ordinal variables (e.g., Likert scales) were compared between groups using Mann-Whitney U tests. Categorical variables were compared between groups using χ^2^ contingency tests. For the subgroup analysis where the N per group was smaller, multi-level categorical variables were binarised (e.g., employment status was coded as “full-time” or not) to prevent single categories from having too few observations (< 5).

For therapy engagement, we employed linear mixed-effects to analyze continuous outcome measures (number of opens, duration in minutes) over time (week) and between groups (intervention or control), taking into account control variables identified in the baseline characteristics comparison between groups (an example model formulation: outcome ∼ group × week + control + (1|participant)). Due to instances where participants may have left the PDF open on their computer or left their phone open on the app, we removed duration outliers ≥ 3 standard deviations from the mean per group (≥ 148 minutes for PDF viewing sessions and ≥ 93 minutes for app sessions).

For symptom reduction scales (primary: GAD-7, PHQ-9; additional: MSQ, WSAS), we first removed participants whose overall change (in any of the four scales) from baseline to their last recorded week exceeded 3 standard deviations from the mean per scale (N = 5 from intervention group, N = 4 from control group). We examined symptom reduction over time using a similar linear mixed-effects modeling approach to the above engagement analysis.

## Results

### Participant recruitment and retention

We screened a total of 2,146 individuals for eligibility in our study, of whom 682 participants (31.78%) met the inclusion criteria (see **<u>Participants</u>** section for inclusion/exclusion criteria and **Fig. 2**). Recruitment for the baseline survey was capped at 540 participants. These participants were randomly assigned to one of two groups: 322 participants to the intervention group and 218 participants to the control group. In the final survey of the study, we identified 12 participants in the control group who reported that they had been using the Limbic app despite not having been instructed to do so (the app was freely available on the app stores). To preserve the integrity of the control condition and ensure that any observed effects could be attributed to the intervention itself, we excluded these 12 participants from further analysis. This adjustment resulted in a final control group of 206 participants.

**Figure 1.**
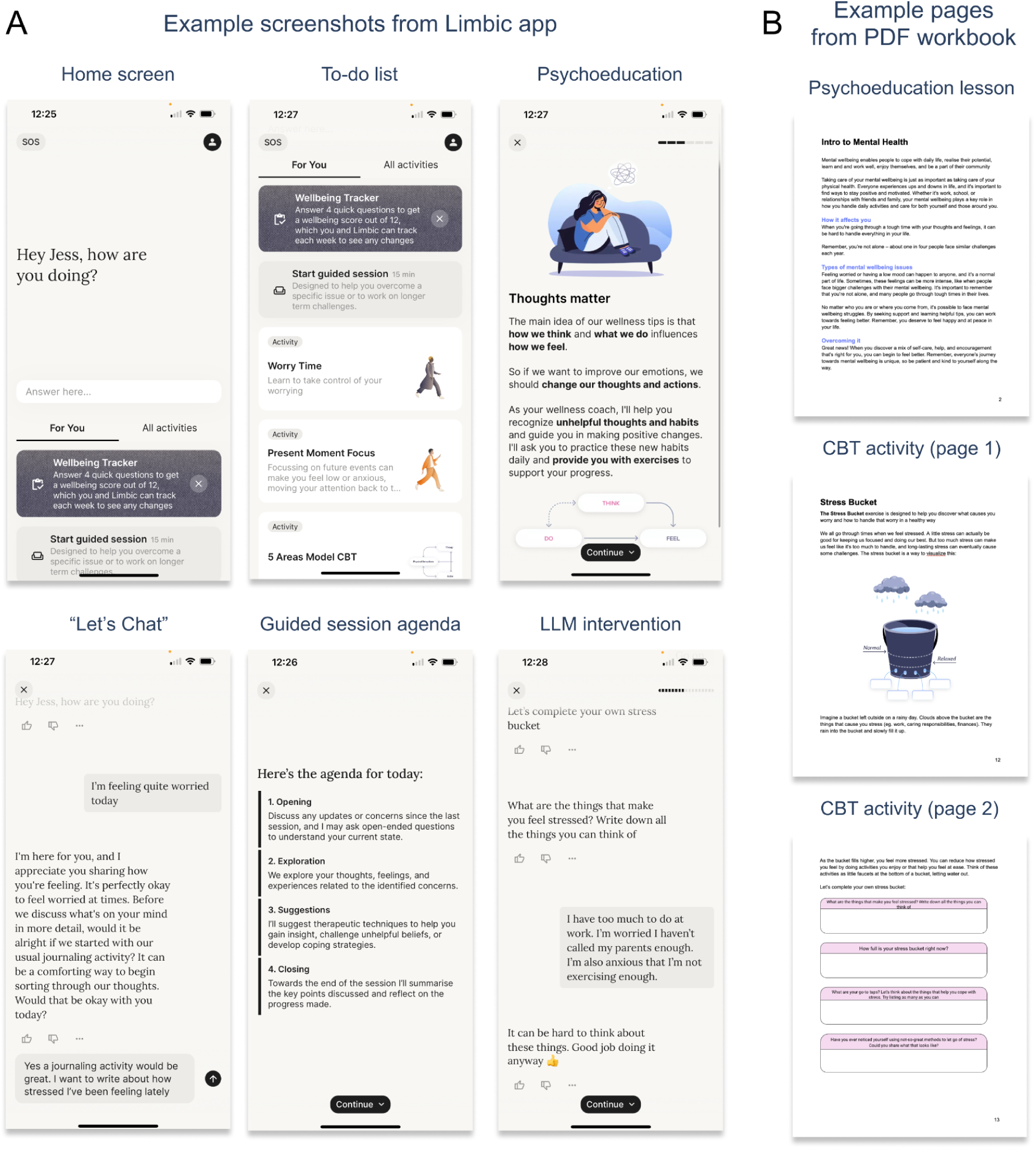
Wellbeing materials provided for the intervention (app) and control (PDF workbook) groups. **(A)** Example screenshots from the Limbic app, featuring (from left to right) the home screen with the “Let’s Chat” feature at the top, followed by the “to-do list” of available psychoeducation lessons and CBT activities, with examples for a Let’s Chat conversation, guided session agenda, and CBT activity (or “intervention”) presented via a conversational interface using an LLM. **(B)** Example pages from the PDF workbook equivalent to the Limbic app, with psychoeducation lessons and CBT activities presented as text and images.

**Figure 2.**
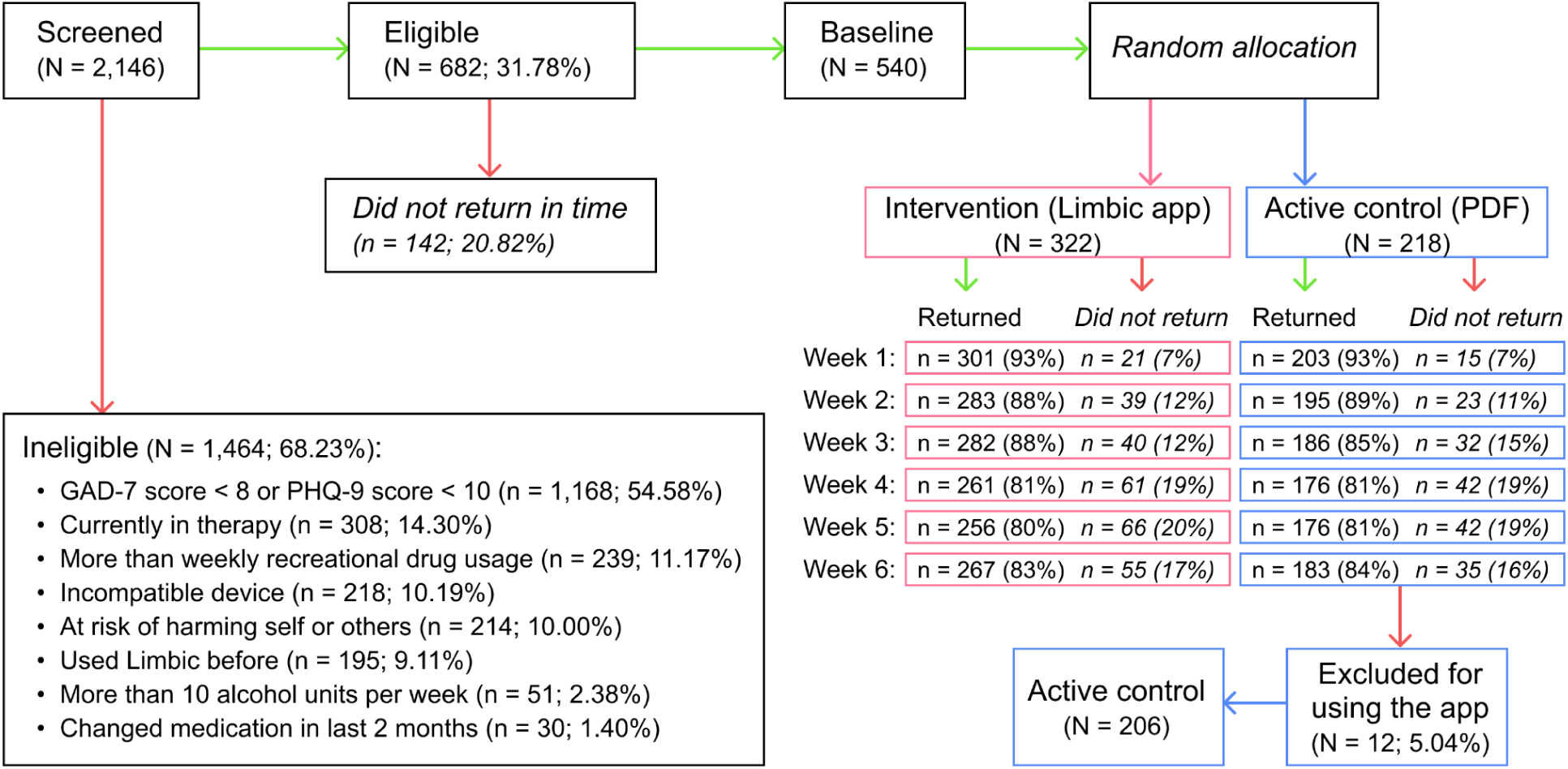
CONSORT diagram for the randomized controlled trial (RCT). GAD-7 is the Generalized Anxiety Disorder 7-item scale and PHQ-9 is the Patient Health Questionnaire 9-item scale.

To assess the equivalence of the groups at baseline, we compared demographic and background characteristics between the intervention and control groups. We found that the intervention group had a slightly lower average score on the GAD-7 compared to the control group (mean difference Δ = 1.10, p = 0.004), indicating slightly less anxiety symptoms at baseline. Conversely, the intervention group had a slightly higher average score on the Patient Health Questionnaire 9-item scale (PHQ-9) (Δ = 0.91, p = 0.048), suggesting slightly more depressive symptoms. Additionally, participants in the intervention group reported feeling less comfortable using digital tools (Δ = 0.12 on a 1–5 scale, p = 0.034). All other baseline characteristics, including age, gender, and other clinical measures, did not differ significantly between the groups (see **Supplementary Table 1**).

Throughout the six-week study period, participants from both groups were invited to complete a weekly survey to monitor their progress and engagement. The average weekly survey completion rates were high and comparable between the groups, reflecting strong participant retention: 85.40% ± 4.76% for the intervention group and 84.87% ± 4.60% for the control group. By the final week (Week 6), retention rates remained consistent, with 82.92% of the intervention group and 83.01% of the control group completing the survey.

Given these observed baseline differences in anxiety (GAD-7), depression (PHQ-9), and how comfortable participants felt with using digital tools, we controlled for these variables in all subsequent analyses.

### Enhanced therapy engagement for app than PDF

#### USAGE FREQUENCY

First, we compared how frequently participants engaged with the therapeutic materials by examining the number of times the PDF workbook was opened in the control group versus the app in the intervention group (see **Fig. 3A**). Control participants opened the PDF an average of 3.9 ± 5.0 times over the six-week period, with the number of opens per week declining over time—from 1.8 ± 1.5 times in Week 1 down to 0.2 ± 0.7 times in Week 6. This decline suggests waning engagement with the PDF workbook as the study progressed.

**Figure 3.**
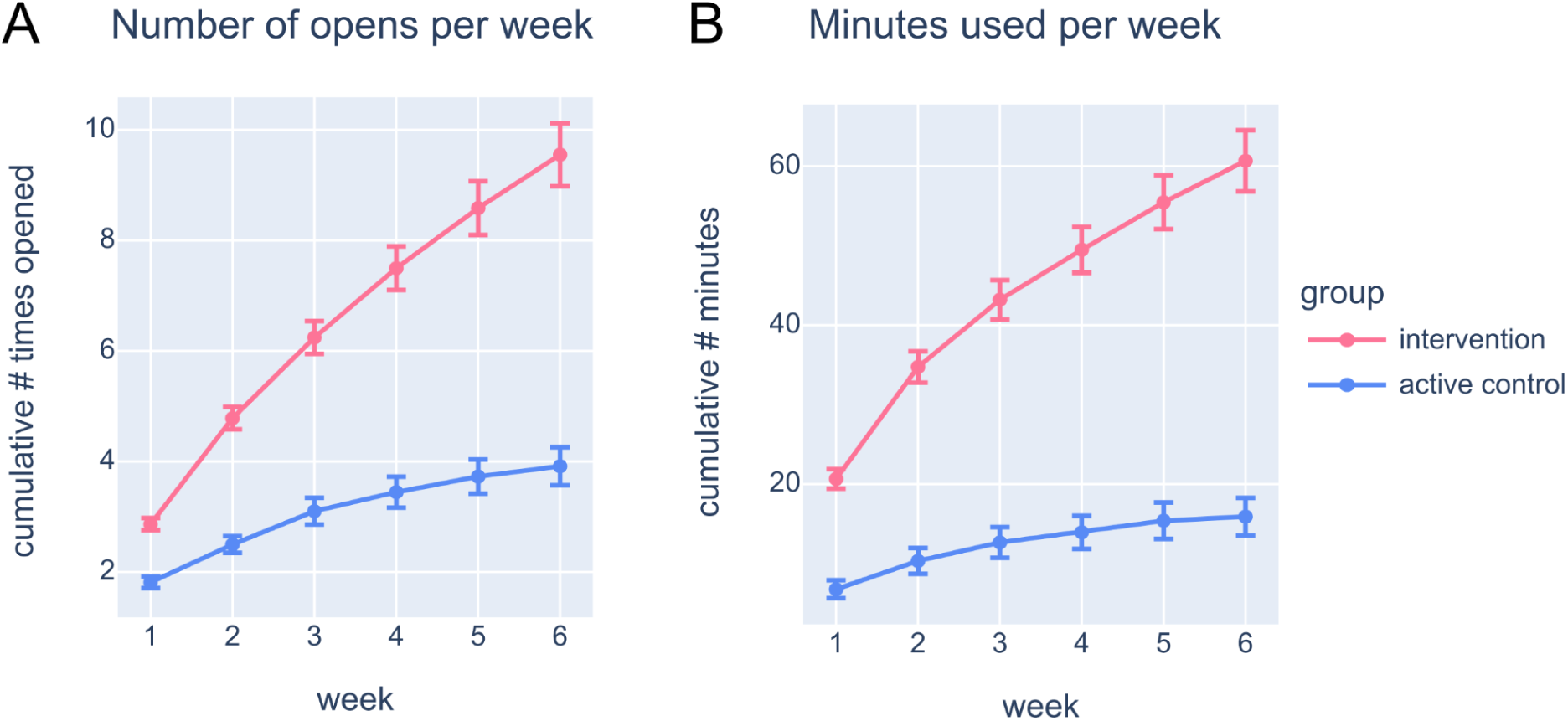
Increased engagement with the Limbic App. **(A)** Cumulative number of opens automatically logged by the app (intervention: pink) or PDF workbook (control: blue) per week, with the mean shown per group. Error bars indicate standard error. **(B)** Cumulative usage duration (in minutes) of the app (intervention) or PDF workbook (control) per week, with the mean shown per group.

In contrast, intervention participants opened the app a total of 9.6 ± 10.2 times over six weeks, more than double (2.4 times) the frequency of the control group. Similar to the control group, the number of app opens per week decreased over time, from 2.9 ± 2.0 times in Week 1 (1.6 times higher than the control group) to 1.0 ± 1.8 times in Week 6 (5 times higher than the control group). This overall higher usage frequency for the intervention group indicates that the app maintained better engagement throughout the study than the PDF workbook (main effect of group: β = −1.207, p < 0.001), an effect that amplified throughout the study period. This suggests that the interactive and personalized nature of the app effectively encouraged participants to engage with the therapeutic content more frequently than the static PDF workbook and enables sustained engagement over an extended period.

#### USAGE DURATION

We also assessed the total duration participants spent engaging with the therapeutic materials (see **Fig. 3B**). Control participants viewed the online PDF for a total of 15.9 ± 34.0 minutes over the six weeks. The longest viewing time occurred in Week 1 (6.8 ± 16.4 minutes), declining sharply to just 0.5 ± 2.5 minutes by Week 6. This trend mirrors the decrease in usage frequency and indicates diminishing engagement with the PDF over time.

In contrast, intervention participants spent a total of 60.7 ± 69.0 minutes using the app, significantly longer than the control group (main effect of group: β = −14.043, p < 0.001). This represents a 3.8-fold increase in total usage duration compared to the control group. Similar to the control group, app usage was highest in Week 1 (20.6 ± 22.1 minutes, 3 times higher than control) and decreased over time to 5.2 ± 12.0 minutes in Week 6 (10.4 times higher than control). The app therefore consistently maintained higher engagement levels each week (main effect of group: β = −14.043, p < 0.001), with app usage duration becoming increasingly magnitudes greater than PDF usage duration throughout the study period. The substantial difference in usage duration suggests that the app not only encouraged more frequent engagement but also sustained longer interactions per session. This sustained engagement is critical, as longer interaction times may enhance the assimilation of therapeutic content and contribute to better clinical outcomes.

### Personalized guided sessions are associated with enhanced well-being

#### OVERALL GROUP COMPARISON

Our other primary outcome measures focused on symptom reduction, specifically targeting anxiety symptoms measured by the Generalized Anxiety Disorder 7-item scale (GAD-7) and depression symptoms measured by the Patient Health Questionnaire 9-item scale (PHQ-9). When considering each group as a whole, without accounting for individual engagement levels, both groups exhibited reductions in anxiety and depression symptoms over the six-week period (main effect of week on GAD-7: β = −0.422, p < 0.001; and on PHQ-9: β = −0.441, p < 0.001; see **Fig. 4A**). The intervention group showed a mean reduction (from baseline to the final measured week per participant) of −2.70 ± 3.98 on the GAD-7 and −2.97 ± 4.22 on the PHQ-9. Similarly, the control group demonstrated a mean reduction of −3.46 ± 4.19 on the GAD-7 and −2.81 ± 4.77 on the PHQ-9. Across all weeks, both groups had comparable GAD-7 scores (main effect of group: β = −0.037, p = 0.882) and PHQ-9 scores (main effect of group: (β = −0.113, p = 0.687). The rate of decline in symptom severity was also equivalent between groups for GAD-7 (group × week: β = −0.031, p = 0.511) and PHQ-9 (group × week: β = 0.050, p = 0.328). This indicates that, on average, the Limbic app had a comparable clinical effect to standard care when disregarding individual engagement levels.

**Figure 4.**
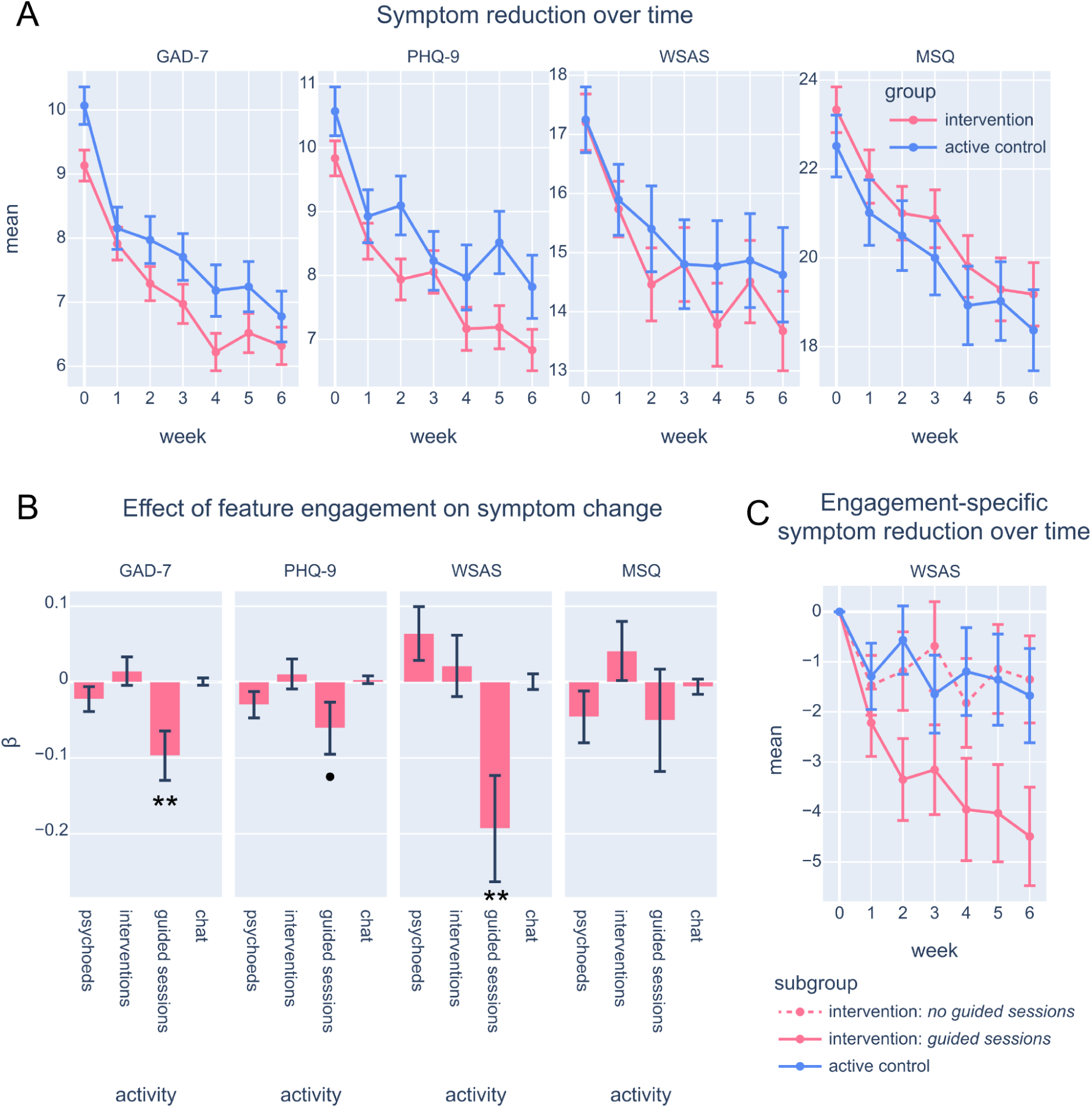
Symptom reduction over time. **(A)** Generalized Anxiety Disorder score (GAD-7), Patient Health Questionnaire (PHQ-9) score, Work and Social Adjustment Scale (WSAS) score, and Mini Sleep Questionnaire (MSQ) score each week (x axis), with the average shown per group (intervention: pink, control: blue). Error bars indicate standard error. **(B)** Beta (β) coefficients for the effect of feature engagement (number of psychoeducation lessons or “psychoeds”, CBT activities or “interventions”, guided sessions, or messages sent in “let’s chat”) on the change in symptom scores over time, taken from separate linear mixed-effects models per measure (GAD-7, PHQ-9, WSAS, and MSQ). Error bars indicate standard error. ** p < 0.01, • p < 0.10 **(C)** Same as the WSAS panel in **A**, except shown for participants in the control group (blue) with sufficient activity (at least 5 minutes of viewing, and viewed at least 6 pages), participants in the intervention group who did guided sessions (pink), and participants in the intervention group who did not do guided sessions but did at least one psychoeducation lesson or CBT activity (pink, dashed). ** p < 0.01, • p < 0.10, from linear mixed effects model predicting symptom score (x axis, as separate models) by a group (control, guided sessions, other activities) × time (week) interaction.

We also assessed additional clinically relevant measures: the Work and Social Adjustment Scale (WSAS) for general well-being and the Mini Sleep Questionnaire (MSQ) for sleep disorder symptoms. Consistent with the GAD-7 and PHQ-9, there were no significant differences between the intervention and control groups on the WSAS (main effect of group: β = −0.649, p = 0.326; group × week effect: β = 0.053, p = 0.587). For the MSQ, although the control group had lower scores across all weeks (including baseline), the group × week interaction was not significant (main effect of group on MSQ: β = −1.688, p = 0.024; group × week effect on MSQ: β = 0.041, p = 0.678) which suggests that sleep symptoms improved at a similar rate for both groups

#### IMPACT OF INTERVENTION ENGAGEMENT ON SYMPTOM REDUCTION

Recognizing that overall group comparisons might mask the effects of individual engagement with specific app features, we investigated whether engagement with certain features predicted different degrees of symptom reduction. A key feature of the genAI-enabled app is the **“**guided sessions,**“** which provide clinical personalization achievable only through the integration of domain-specific AI tailored to mental health. This feature allows users to explore specific issues or challenges they are experiencing through guided questioning. The chatbot – enhanced by specialized clinical machine learning algorithms designed explicitly for mental health applications – identifies core psychological mechanisms underlying the user’s problems. By augmenting the generative AI with this domain-specific clinical AI, the app recommends tailored CBT exercises that directly address the identified issues, providing a uniquely personalized therapeutic experience.

Among the 322 participants in the intervention group, engagement with app features was high but somewhat varied: 66.77% started at least one CBT activity, 68.94% completed at least one psychoeducation lesson, 81.99% sent at least one message using the ‘Let’s Chat’ feature, and 29.81% completed at least one guided session. The average completion time for those who started a CBT activity was 4.3 minutes (± 8.4 minutes). Participants who sent two or more messages had a conversation for 6.7 minutes on average (± 9.4 minutes). Guided sessions were the longest activities, with participants taking on average 11.2 minutes (± 7.0 minutes) to complete them. Note that completion time data for psychoeducation lessons was not collected due to a technical error.

We included the amount of engagement with each of these four features in our statistical models to assess their impact on symptom reduction (GAD-7, PHQ-9, MSQ, and WSAS). Our analysis revealed that participants who engaged more with the guided sessions showed a significantly greater improvement in anxiety symptoms over time, as measured by GAD-7 scores (guided sessions × week interaction: β = −0.097, p = 0.003; see **Fig. 4B**). There was also a trend toward greater improvement in depression symptoms (PHQ-9) for those engaging with guided sessions (guided sessions × week: β = −0.060, p = 0.077). Additionally, WSAS scores, reflecting general well-being, reduced most for participants who completed guided sessions (guided sessions × week: β = −0.193, p = 0.006). Engagement with other app features did not show significant associations with symptom reduction, and MSQ scores for sleep disorder severity were not significantly influenced by engagement levels with any features (all p > 0.379). These findings suggest that the personalized, clinically tailored experience provided by the guided sessions is a key driver of symptom improvement within the intervention group, highlighting the importance of domain-specific genAI in enhancing therapeutic outcomes.

#### ENGAGEMENT SUBGROUPS ACROSS INTERVENTION AND CONTROL

To further elucidate the impact of guided sessions, we conducted an exploratory analysis by grouping participants based on their engagement levels. Within the intervention group, we identified two subgroups: 1) intervention participants who completed at least one guided session (n = 94), and 2) intervention participants who did not engage with guided sessions but started at least one CBT activity and completed one psychoeducation lesson (n = 103). For a fair comparison, we selected a control subgroup of participants who viewed the PDF workbook for at least five minutes and viewed at least six pages, which was the point at which the first CBT activity appears in each course (n = 90). We also assessed whether these three subgroups differed on any baseline characteristics, and controlled for significant differences in gender distribution (χ^2^ = 8.048, p = 0.018), full-time employment (χ^2^ = 6.073, p = 0.048), having had a previous diagnosis (χ^2^ = 7.025, p = 0.030), and Obsessive-Compulsive Index Revised (OCI-R) scores (t = 2.912, p = 0.004) in subsequent analyses.

We compared these roughly equal-sized groups on our primary clinical outcome measures (GAD-7 and PHQ-9) and additional measures (WSAS and MSQ) using a linear mixed-effects model. The analysis showed that the guided session group did not significantly modulate symptom reduction for GAD-7 (all p > 0.210), PHQ-9 (all p > 0.460), or MSQ (all p > 0.539; see **Fig. 4C**). However, there was a significantly greater reduction in WSAS scores over time for the subgroup that completed guided sessions compared to the control group (guided sessions group × week: β = −0.316, p = 0.033). Importantly, this effect was significant even after accounting for differences in the total duration of app or PDF usage by each participant. This suggests that it was not merely the amount of time spent engaging with the materials but the qualitative aspect of the guided sessions feature that drove the significant improvement in general well-being. Therefore, the clinical personalization of guided sessions appears to enhance the therapeutic experience beyond what is achievable through standard CBT activities or psychoeducation lessons alone, even when digitized.

### Comparable safety of intervention and control groups

Ensuring the safety of participants is crucial when introducing new therapeutic interventions, especially those involving emerging technologies like generative AI. Throughout the study period, we closely monitored the occurrence of adverse health events by asking participants each week whether they had experienced any new adverse events. These could include any negative changes in physical health (e.g., falling and hurting themselves) or mental health (e.g., experiencing stress at work), regardless of whether they were directly related to the intervention. All incidents were monitored on a weekly basis by clinical researchers so that any severe events would be detected and the participant referred to crisis support.

After controlling for baseline anxiety and depression levels (GAD-7 and PHQ-9 scores), we assessed the likelihood of participants reporting any adverse event during the study. The proportion of participants who ever reported an adverse event was nearly identical between the two groups: 36.96% in the intervention group and 38.22% in the control group (β = 0.002, p = 0.959). Thus, participants using the genAI-enabled app were no more likely to experience adverse events than those experiencing standard care.

We also examined the total number of adverse events reported by each group to assess any differences in the overall safety profile. Fewer adverse events were reported per participant in the intervention group (0.59 per participant, with a total of 190 events) than in the control group (0.71 per participant, with a total of 149 events), although this difference was not significant (β = 0.093, p = 0.310).

Importantly, there were no serious adverse events reported in either group that were directly related to the intervention. The absence of serious adverse events and the comparable rates of overall adverse events between the groups indicate that the genAI-enabled Limbic app was as safe as the active control over the 6 week study period.

### Enhanced usability and satisfaction of app compared to PDF

Throughout the study period, we surveyed participants in both groups weekly to gather their impressions of the tool they had been assigned—the genAI-enabled app for the intervention group or the PDF workbook for the control group (see **<u>Supplementary materials</u>** for list of questionnaire items). We focused on key aspects such as usability, satisfaction, and perceived learning to evaluate the user experience comprehensively.

When averaging these additional measures across all six weeks, we found that participants in the guided sessions subgroup reported significantly higher usability scores compared to both the control subgroup and the intervention subgroup that did not engage with guided sessions (see **Fig. 5A**). Specifically, the guided sessions subgroup found the app more accessible than the control subgroup (β = −0.545, p < 0.001, p_FDR_ = 0.016) and also found it easier to navigate (β = −0.474, p < 0.001, p_FDR_ = 0.002). These results indicate that usability was significantly associated with the use of guided sessions, suggesting that this feature enhances the overall accessibility and user-friendliness of the app.

**Figure 5.**
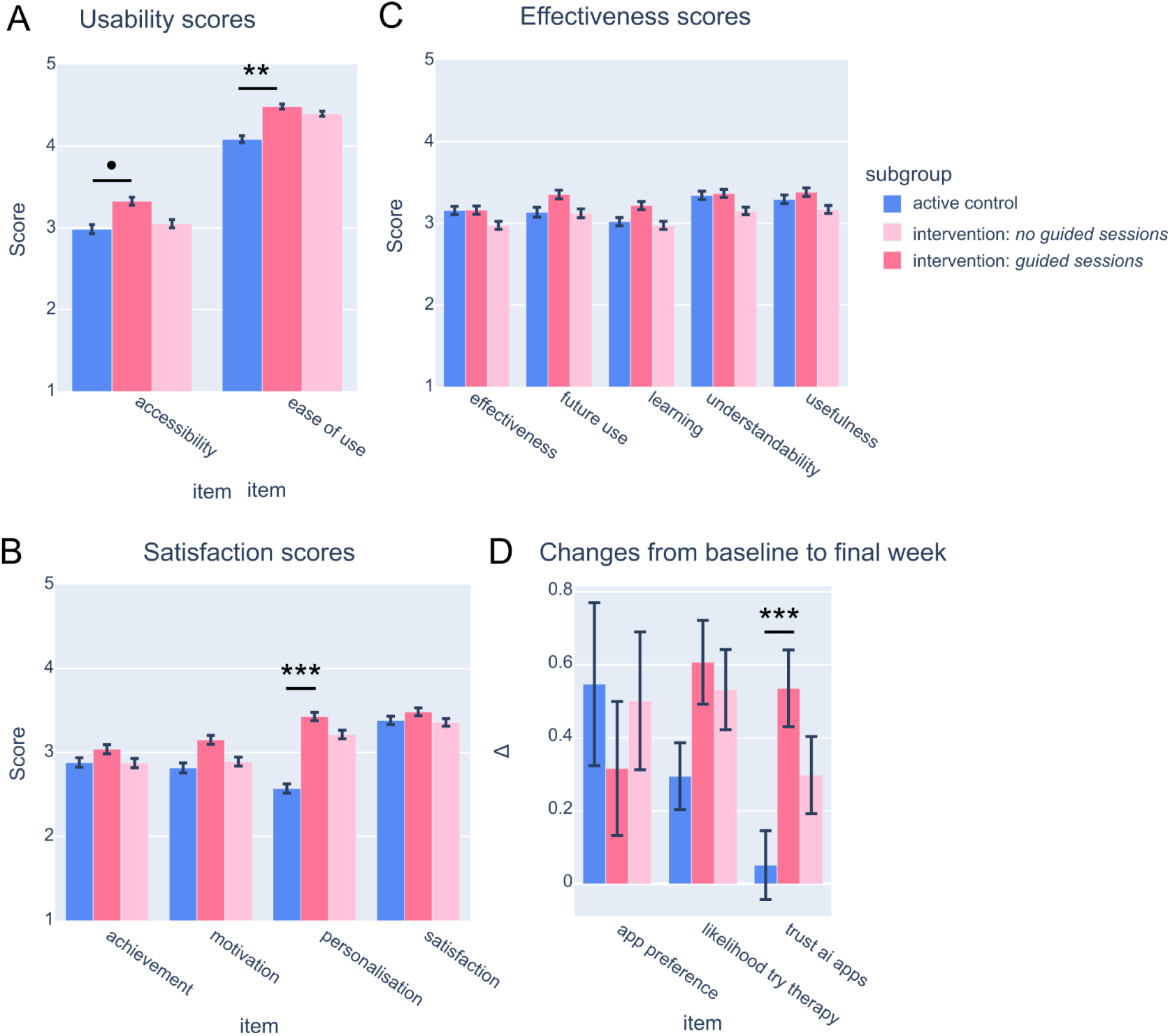
Ratings of effectiveness, satisfaction, and usability. **(A)** Average rating for each usability survey item across the 6 weekly surveys per subgroup (control, n=90: blue; intervention without guided sessions, n=103: light pink; intervention with guided sessions, n=94: pink). Error bars represent standard error. **(B)** Same as **A** but for satisfaction items. *** p < 0.001 **(C)** Same as **A** but for effectiveness items. **(D)** Change from baseline to the final week 6 survey.

Self-reported satisfaction levels were higher in the guided sessions subgroup across multiple dimensions (see **Fig. 5B**). Participants who engaged with guided sessions reported greater feelings of achievement, higher motivation to use the tool, increased overall satisfaction, perceived the course as more structured, expressed a greater willingness to continue using the tool in the future, and felt that the tool was more personalized to their needs. Among these, the most significant difference was observed in the perception of personalization, which was significantly higher in the guided session subgroup than in the control subgroup (β = −0.987, p < 0.001, p_FDR_ < 0.001). This suggests that the personalized experience provided by the guided sessions significantly enhances user satisfaction and may contribute to sustained engagement.

Regarding self-reported learning outcomes, the guided sessions subgroup also reported higher levels compared to the control group (see **Fig. 5C**). They felt that the tool was more useful, learned more from it, were better able to translate learnings into everyday life, understood the content better, and found the tool overall more beneficial. These differences, however, were not statistically significant after correcting for multiple comparisons (all p_FDR_ > 0.129).

Finally, we examined how key subjective measures changed from baseline to the final week of the study across the different subgroups. All subgroups reported an increase in their trust in well-being apps that use AI over the six-week period (main effect of week: β = −0.537, p < 0.001; see **Fig. 5D**). However, this increase was most pronounced in the guided sessions subgroup compared to the control group (week × subgroup interaction: β = −0.474, p = 0.001, p_FDR_ = 0.007). Similarly, the guided sessions subgroup showed a larger increase in their likelihood of seeking a therapy session with a human therapist in the next three months compared to the control group, although this was not significantly larger after correcting for multiple comparisons (week × subgroup: β = −0.307, p = 0.043, p_FDR_ = 0.724). There were no significant differences between the subgroups (all p > 0.587) on the change in preference for apps vs digital workbooks, as all participants showed a preference for apps at both time points.

## Discussion

In this randomized controlled trial, we compared a generative AI-enabled therapy delivery tool to digital workbooks delivering static CBT content, akin to the current standard of care for before and between therapy sessions. We found that the intervention group exhibited a threefold increase in engagement with therapeutic activities compared to the control group. This suggests that delivering CBT content through generative AI substantially enhances patient engagement in mental health treatment. Furthermore, the genAI-enabled therapy tool was as safe as the active control, with no indication of increased adverse events, indicating that generative AI can be safely used in clinical contexts when combined with appropriate safety guardrails. Finally, our investigation into the efficacy of the genAI-enabled tool revealed that engagement with Limbic’s clinical personalization feature was associated with greater symptom reduction over the 6-week study period compared to the active control group. These results suggest that generative AI can significantly impact patient outcomes in terms of engagement, safety, and symptom reduction, especially when utilizing clinical personalization to tailor exercises to the user’s specific problems.

A significant hurdle in CBT is encouraging patients to consistently engage with therapeutic homework, which is essential for the treatment’s effectiveness (Malik et al., 2022; Oewel et al., 2024; Torous et al., 2018). Our study tackles this pervasive problem by demonstrating that delivering CBT activities through an interactive, genAI-enabled app markedly enhances patient engagement compared to conventional digital workbooks. Specifically, the app was approximately three times more engaging, with 2.4 times higher open rates and 3.8 times longer usage durations. Interestingly, these engagement benefits magnified over the study period, suggesting that the usage of generative AI is especially advantageous for the engagement of patients over longer periods of times (weeks or months), as often encountered on waitlists for psychotherapy. This elevated engagement was also evident in subjective ratings, where the intervention group reported significantly higher scores on ease of use and personalization than the control group. These findings are in line with previous research finding that people generally hold positive attitudes towards chatbot-delivered CBT (Limpanopparat et al., 2024). Other studies have also reported increased engagement with chatbots compared to traditional static materials (Yasukawa et al., 2024), although in some cases the enhanced engagement with the Limbic app is substantially greater than that reported in other studies (Karkosz et al., 2024; Perski et al., 2019), which could be attributable to Limbic’s proprietary clinical AI. Together, this suggests that leveraging generative AI to create interactive and personalized digital materials can effectively overcome the longstanding challenge of low engagement in CBT homework, providing a more compelling and user-friendly platform that increases touchpoints between patients and their therapy programs.

A persistent challenge in mental health care is not just delivering therapeutic content but encouraging meaningful patient engagement that translates into symptom reduction (Conklin & Strunk, 2015; Sapkota et al., 2023). In our study, we observed that while both the intervention group using the genAI-enabled app and the control group using a PDF workbook demonstrated similar rates of improvement in anxiety (GAD-7), depression (PHQ-9), sleep disturbances (MSQ), and general well-being (WSAS), the app-based delivery proved more engaging overall. These results indicate that app-based interventions can be as clinically effective as traditional methods – as is typically observed in RCTs that use active control conditions (Moshe et al., 2021) – while offering enhanced user engagement, aligning with results reported by similar studies comparing digital solutions to traditional, static CBT material (Jonassaint et al., 2024; Quero et al., 2019). However, our further analyses unmasked an important role of engagement patterns in improving clinical outcomes.

A closer examination of engagement patterns showed that participants who frequently used the genAI-driven “guided sessions” feature, designed to closely replicate a patient-therapist interaction, experienced significantly greater reductions in anxiety symptoms and improvements in well-being compared to those who engaged primarily with psychoeducation lessons, CBT activities, or the “Let’s Chat” feature. Subgroup analyses also underscored a distinct advantage in well-being over time for the guided sessions users compared to other app users and the control group. Crucially, these benefits persisted after controlling for overall app usage time and baseline symptom levels, suggesting that the clinical personalization offered by the genAI-guided sessions played a central role in symptom improvement. This finding supports previous evidence for the personalization of CBT interventions improving therapy adherence (Cheung et al., 2018; Fitzsimmons-Craft et al., 2024) and for engagement with LLM-delivered CBT content improving mental health (Bhatt, 2024; Kim et al., 2024). As an example, one RCT observed a larger decrease in loneliness for users who engaged with a CBT chatbot compared to those who did not engage, or to those who engaged with a self-help book, despite total engagement time being similar between these subgroups (Karkosz et al., 2024). Overall, these findings underscore the importance of engagement with personalized, clinically tailored content powered by genAI, suggesting that such personalization is essential for optimizing both clinical outcomes and patient engagement in mental health interventions.

Applying this personalized, genAI-driven approach across various contexts — such as supporting individuals who cannot access therapy, offering preventative care for those with mild symptoms without a human therapist, assisting patients between therapy sessions, or aiding individuals on therapy waitlists — could lead to patients recovering before beginning treatment or starting human-led therapy in a better state (Beg et al., 2024; Sadeh-Sharvit et al., 2023). By reducing symptoms and enhancing preparation through personalized interventions, patients are less likely to deteriorate while waiting for treatment to begin, ultimately improving the efficiency and effectiveness of mental health services (Anderson et al., 2019; Rollwage, Habicht, Juechems, et al., 2024).

The enhanced clinical outcomes observed in participants who engaged with the personalized, genAI-driven features highlight the critical role of clinical personalization in digital mental health interventions. However, achieving this level of personalization is not attainable using large language models (LLMs) in isolation. While LLMs excel at generating human-like text and facilitating interactive conversations, they lack the specialized clinical knowledge and adherence to therapeutic protocols necessary for safe and effective mental health support (Abd-Alrazaq et al., 2020; Li et al., 2023). To bridge this gap, the Limbic app employs a combined architecture that integrates LLMs with specialized clinical machine learning models tailored explicitly for mental health applications. These domain-specific models are designed to identify and understand core psychological mechanisms underlying a user’s problems by analyzing inputs through a clinically informed lens. By constraining and guiding the behavior of the LLM with these specialized models, the app ensures that all interactions align with established clinical guidelines and therapeutic best practices.

In line with this, the findings of this RCT suggest that the genAI-enabled Limbic app is as safe as the standard PDF workbooks used in typical care settings. The comparable safety profiles between the intervention and control groups address a critical concern regarding the use of AI technologies in mental health interventions – that is, the potential for unpredictable or harmful responses that could damage a user’s mental or physical health. While large providers of LLMs (e.g., OpenAI) invest substantially in generic safety guardrails, the mental health context is particularly sensitive and requires significantly enhanced domain-specific safety measures. Therefore, the Limbic app was equipped with multiple proprietary safety layers to screen user input and LLM output, detecting any safety-relevant situations and handling them accordingly. Thus, this study demonstrates that highly specific, proprietary safety layers can ensure the safe and effective utilization of generative AI in mental healthcare.

Random allocation of participants to the intervention and an active control, equivalent to standard care, allows us to draw causal conclusions about the impact of genAI-enabled therapy delivery on engagement and treatment outcomes. One limitation, however, is that our evidence for increased well-being among those who engaged with guided sessions was observational rather than experimentally manipulated. Unmeasured factors, such as participants’ inherent motivation levels, could have contributed to the observed improvements in clinical outcomes. Future research can methodically investigate the impact of these genAI-enabled clinical features on engagement and clinical outcomes by randomly assigning participants to have or not have access to different features.

In summary, this RCT has demonstrated that genAI-enabled therapy support has a significant, beneficial effect on increasing engagement with therapeutic exercises and materials. Moreover, the use of this tool was safe and effective when benchmarked against an active control representing standard care. Importantly, engagement with features that provide clinical personalization, made possible with genAI, was associated with significantly improved symptom reduction and enhanced well-being. These results suggest that clinical AI is an effective, scalable solution for improving patient experience and clinical outcomes, from the point of seeking help through to recovery.

## Data Availability

All data produced in the present study are available upon reasonable request to the authors.

## Acknowledgements

This research was funded by Limbic Limited. We would like to thank the research team at Limbic, particularly Sashank Pisupati, George Prichard, Keno Juchems, and Annamari Balogh, for their significant contributions to the development of the clinical AI used in this study. We also thank the wider Limbic team for their support throughout this work.

