## Supplementary Materials for "AI-enabled conversational agent increases engagement with cognitive-behavioral therapy: A randomized controlled trial"

### Tables

| Variable | Intervention | Control | Statistic |
|---|---|---|---|
| GAD-7 | M = 9.15<br>SD = 4.32 | M = 10.25<br>SD = 4.22 | t = -2.882<br>p = 0.004** |
| PHQ-9 | M = 9.87<br>SD = 4.87 | M = 10.78<br>SD = 5.50 | t = -1.983<br>p = 0.048* |
| WSAS | M = 17.26<br>SD = 17.26 | M = 17.39<br>SD = 17.39 | t = -0.17<br>p = 0.87 |
| MSQ | M = 23.52<br>SD = 23.52 | M = 22.59<br>SD = 22.59 | t = 1.10<br>p = 0.27 |
| Familiarity with wellbeing apps | M = 2.28<br>SD = 2.28 | M = 2.25<br>SD = 2.25 | U = 34308.0<br>p = 0.48 |
| Trust in wellbeing apps that use AI | M = 2.51<br>SD = 2.51 | M = 2.52<br>SD = 2.52 | U = 33025.0<br>p = 0.93 |
| Familiarity with CBT | M = 2.19<br>SD = 2.19 | M = 2.24<br>SD = 2.24 | U = 32450.5<br>p = 0.66 |
| Likelihood of trying therapy soon | M = 1.43<br>SD = 1.43 | M = 1.37<br>SD = 1.37 | U = 35224.5<br>p = 0.15 |
| Preference for PDFs over apps for wellbeing | M = 2.45<br>SD = 2.45 | M = 2.50<br>SD = 2.50 | U = 32975.0<br>p = 0.91 |
| Age | M = 37.56<br>SD = 37.56 | M = 35.78<br>SD = 35.78 | t = 1.72<br>p = 0.09 |
| Comfort with digital tools | M = 4.55<br>SD = 4.55 | M = 4.67<br>SD = 4.67 | U = 30674.0<br>p = 0.03* |
| Wellbeing knowledge | M = 2.48<br>SD = 2.48 | M = 2.45<br>SD = 2.45 | U = 33562.0<br>p = 0.81 |
| HAI | M = 19.44<br>SD = 19.44 | M = 19.79<br>SD = 19.79 | t = -0.47<br>p = 0.64 |
| OCI-R | M = 17.28<br>SD = 17.28 | M = 19.09<br>SD = 19.09 | t = -1.65<br>p = 0.10 |
| PCL-5 | M = 25.12<br>SD = 25.12 | M = 26.83<br>SD = 26.83 | t = -1.25<br>p = 0.21 |
| PDSS | M = 3.83<br>SD = 3.83 | M = 3.89<br>SD = 3.89 | t = -0.13<br>p = 0.90 |
| Phobia | M = 8.27<br>SD = 8.27 | M = 7.68<br>SD = 7.68 | t = 0.75<br>p = 0.45 |
| SPIN | M = 26.41<br>SD = 26.41 | M = 26.73<br>SD = 26.73 | t = -0.25<br>p = 0.81 |
| Alcohol consumption | M = 1.10<br>SD = 1.10 | M = 1.14<br>SD = 1.14 | U = 31981.00<br>p = 0.26 |

| | | | |
|---|---|---|---|
| Used wellbeing apps before | Yes = 108 (33.54%) | Yes = 72 (33.03%) | $\chi^2$ = < 0.001<br>p = 0.975 |
| Previously had therapy | Yes = 177 (54.97%) | Yes = 107 (49.08%) | $\chi^2$ = 1.578<br>p = 0.209 |
| Taking psychoactive medication | Yes = 73 (22.67%) | Yes = 45 (20.64%) | $\chi^2$ = 0.206<br>p = 0.650 |
| Chosen course for study | Low mood = 94 (29.19%)<br>Sleep = 62 (19.25%)<br>Worry = 147 (45.65%)<br>Not selected = 19 (5.90%) | Low mood = 67 (30.73%)<br>Sleep = 52 (23.85%)<br>Worry = 99 (45.41%)<br>Not selected = 0 (0%) | $\chi^2$ = 0.928<br>p = 0.629 |
| Gender | Female = 224 (69.57%)<br>Male = 91 (28.26%)<br>Non-binary = 6 (1.86%)<br>Other = 1 (0.31%) | Female = 160 (73.39%)<br>Male = 51 (23.39%)<br>Non-binary = 7 (3.21%)<br>Other = 0 (0%) | $\chi^2$ = 2.417<br>p = 0.299 |
| Ethnicity simplified | Asian = 27 (8.39%)<br>Black = 27 (8.39%)<br>Mixed = 30 (9.32%)<br>Not stated = 2 (0.62%)<br>Other = 15 (4.66%)<br>White = 221 (68.63%) | Asian = 20 (9.17%)<br>Black = 21 (9.63%)<br>Mixed = 16 (7.34%)<br>Not stated = 1 (0.46%)<br>Other = 11 (5.05%)<br>White = 149 (68.35%) | $\chi^2$ = 0.959<br>p = 0.916 |
| Employment status | Due to start a new job within the next month = 3 (0.93%)<br>Full-Time = 166 (51.55%)<br>Not in paid work (e.g. homemaker\', \'retired or disabled) = 32 (9.94%)<br>Not stated = 24 (7.45%)<br>Other = 17 (5.28%)<br>Part-Time = 43 (13.35%)<br>Unemployed (and job seeking) = 37 (11.49%) | Due to start a new job within the next month = 5 (2.29%)<br>Full-Time = 92 (42.20%)<br>Not in paid work (e.g. homemaker\', \'retired or disabled) = 28 (12.84%)<br>Not stated = 15 (6.88%)<br>Other = 5 (2.29%)<br>Part-Time = 38 (17.43%)<br>Unemployed (and job seeking) = 35 (16.06%) | $\chi^2$ = 6.839<br>p = 0.145 |
| Student status | No = 231 (71.74%)<br>Not stated = 34 (10.56%)<br>Yes = 57 (17.70%) | No = 157 (72.02%)<br>Not stated = 19 (8.72%)<br>Yes = 42 (19.27%) | $\chi^2$ = 0.625<br>p = 0.732 |
| Accessibility issues | Vision = 18 (5.59%)<br>Hearing = 14 (4.35%)<br>Mobility = 25 (7.76%)<br>Dexterity = 11 (3.42%)<br>Learning = 25 (7.76%)<br>Memory = 24 (7.45%)<br>Stamina = 29 (9.01%)<br>Social = 42 (13.04%)<br>Other = 9 (2.80%)<br>Prefer not to answer = 5 (1.55%)<br>None = 207 (64.29%) | Vision = 17 (8.25%)<br>Hearing = 5 (2.43%)<br>Mobility = 9 (4.37%)<br>Dexterity = 5 (2.43%)<br>Learning = 21 (10.19%)<br>Memory = 15 (7.28%)<br>Stamina = 24 (11.65%)<br>Social = 40 (19.42%)<br>Other = 1 (0.49%)<br>Prefer not to answer = 2 (0.97%)<br>None = 119 (57.77%) | $\chi^2$ = 11.134<br>p = 0.194 |
| Operating system | iPhone = 232 (72.05%)<br>Android = 89 (27.64%)<br>Not stated = 1 (0.31%) | iPhone = 158 (72.48%)<br>Android = 57 (26.15%)<br>Not stated = 3 (1.38%) | χ2 = 0.044<br>p = 0.833 |

**Supplementary Table 1. Baseline characteristics comparison.** Comparisons between different continuous and categorical variables between the intervention and control groups at baseline. The mean (M) and standard deviation (SD) is shown for continuous variables, compared with independent groups t-tests. The number and proportion of participants included in each category is shown for binary and categorical variables, compared using contingency $\chi^2$ tests across all categories (note that only one response is shown for binary variables).

### Materials

| Category | Label | Question |
|---|---|---|
| **Usability** | accessibility | *How well did the {tool} accommodate your needs and preferences?* |
| | ease of use | *How easy was it to navigate the {tool}?* |
| **Effectiveness** | effectiveness | *How much did the {tool} equip you with the skills to deal with mental health challenges?* |
| | future use | *How likely are you to use the {tool} in the future to improve your mental wellbeing?* |
| | learning | *How much did you apply the information learnt from the {tool} in your daily life?* |
| | understandability | *How much did the {tool} help you in understanding and managing your mental health?* |
| | usefulness | *How useful did you find the {tool} for your mental health?* |
| **Satisfaction** | achievement | *How much did the {tool} give you a sense of achievement?* |
| | motivation | *How motivated were you to use the {tool} to improve your mental health?* |
| | personalization | *To what degree did the {tool} provide a personalized experience?* |
| | satisfaction | *Overall, how satisfied are you with the {tool}?* |
| **Change scores** | app preference* | *Imagine that the course you completed in the {tool} was instead offered to you in the form of a {other group tool}. Which format would you prefer?* |
| | likelihood try therapy | *How likely are you to arrange to see a therapist in the next 3 months?* |
| | trust ai apps | *In general, how much do you trust wellbeing apps that use artificial intelligence (AI)?* |

**Supplementary Table 2. Usability and satisfaction questionnaire items.** Items included in the weekly survey, plus the "change score" items that were only included in the baseline survey and the final (week 6) survey. The phrase "{tool}" was replaced by either "digital workbook" for the active control group or "app" for the intervention group. Each item was rated on a Likert scale from 1 to 5. * This item was reverse-scored for the active control group so that higher scores indicate a preference for an app over a digital workbook.
